# Shared Genetic Liability across Systems of Psychiatric and Physical Illness

**DOI:** 10.1101/2024.08.02.24311427

**Authors:** Jeremy M. Lawrence, Isabelle F. Foote, Sophie Breunig, Lukas S. Schaffer, Travis T. Mallard, Andrew D. Grotzinger

## Abstract

Epidemiological literature has shown that there are extensive comorbidity patterns between psychiatric and physical illness. However, our understanding of the multivariate systems of relationships underlying these patterns is poorly understood. Using Genomic SEM and Genomic E-SEM, an extension for genomic exploratory factor analysis that we introduce and validate, we evaluate the extent to which latent genomic factors from eight domains, encompassing 76 physical outcomes across 1.9 million cases, evince genetic overlap with previously identified psychiatric factors. We find that internalizing, neurodevelopmental, and substance use factors are broadly associated with increased genetic risk sharing across all physical illness domains. Conversely, we find that a compulsive factor is protective against circulatory and metabolic illness, whereas genetic risk sharing between physical illness factors and psychotic/thought disorders was limited. Our results reveal pervasive risk sharing between specific groups of psychiatric and physical conditions and call into question the bifurcation of psychiatric and physical conditions.

## Introduction

Individuals with one psychiatric disorder are at such an elevated risk for a second psychiatric diagnosis^1^ that comorbidity is the norm rather than the exception^2^. Critically, this problem extends to physical disorders with multimorbidity (e.g., the presence of ≥ 2 chronic conditions) observed in ∼38% of the global population^3^. The separation of psychiatric from physical diseases reflects a historical dichotomy distinguishing disorders with characterizable physical changes (e.g., organic) from those without (e.g., functional)^4^. However, prior research utilizing large-scale national registries indicates that pervasive comorbidity extends beyond this theoretical distinction, with a psychiatric diagnosis also associated with an increased likelihood of developing a physical illness^5,6^. In isolation, psychiatric conditions contribute substantially to excess mortality^7–9^, diminished health-related quality of life^10^, reduced functional status^11^, and increased primary care burdens^12^. However, the co-occurrence of psychiatric and physical illness exacerbates these pertinent clinical outcomes^10–13^. For example, early mortality amongst diabetes cases is 38% higher when comorbid with a diagnosis of major depression^13^. Given these significant clinical implications, understanding the etiological factors that may underlie these comorbidities is crucial and has been a significant challenge in the field.

Despite the high prevalence of psychiatric and physical comorbidities, understanding the intricate relationships underlying these comorbidities has historically been hindered by practical challenges in obtaining large samples that encompass a variety of psychiatric and physical outcomes. These challenges have limited the application of multivariate approaches capable of identifying shared risk pathways across domains of illness, while also accounting for shared risk within domains. As a result, it has been difficult to obtain a comprehensive overview of the systems of relationships across psychiatric and physical outcomes that reflect the shared etiologies across groupings of traits. Further, even in cases where psychiatric and physical comorbidity between pairs or sets of outcomes is observed, it is ambiguous whether the effect is driven by broader, transdiagnostic risk pathways across multiple conditions. For example, literature investigating patterns of physical comorbidity among patients with depression has found an increased risk for coronary artery disease^14^. However, it remains uncertain if the risk pathways shared with coronary artery disease are unique to major depression or, for example, shared across other internalizing disorders (e.g., anxiety) that phenotypically^1^ and genetically^15^ overlap. The current study leverages cutting-edge multivariate genomic methods to circumvent these pragmatic barriers to examine the shared and unique risk pathways across psychiatric and physical health outcomes.

Genomic Structural Equation Modeling (SEM^16^) extends bivariate frameworks for estimating genetic overlap, such as LD-score regression (LDSC^17^), into a multivariate approach designed to model the genetic overlap of multiple traits. As genome-wide association study (GWAS) summary statistics need not come from the same participant sample, Genomic SEM offers the unique opportunity to examine genetic overlap across a broad range of rare and potentially mutually exclusive outcomes. Here we apply Genomic SEM and introduce and validate a new extension, Genomic Exploratory SEM (E-SEM), to explore the relationships across psychiatric and physical latent genetic factors. This approach allows us to: *(i)* include a broad range of psychiatric and physical health phenotypes ascertained from different participant samples in the same statistical model; (*ii)* conduct exploratory factor analyses to identify genomic factors indexing shared risk across groupings of traits; and *(iii)* delineate between shared psychiatric and physical illness risk pathways that are operating at the broad level of inter-factor correlations or between more circumscribed sets of disorders. Collectively, these results provide a critical context for interpreting an existing phenotypic literature that consistently links psychiatric and physical health outcomes.

## Results

### Factor Structures across Physical Illness Domains

Through a comprehensive analysis of physical illness phenotypes spanning eight domains (neurological, respiratory, circulatory, digestive, endocrine/metabolic, genitourinary, musculoskeletal, and cancer) we identified 76 physical disorders and symptoms across 1,978,608 cases that represent the largest publicly available GWAS summary statistics for each trait. We first examined a common factor structure in each domain. When this simple structure did not provide adequate model fit, we utilized Genomic E-SEM to conduct exploratory factor analyses (EFAs). We validate key features of Genomic E-SEM through a series of simulations and demonstrate its flexibility to identify complex factor structures across a wide range of physical outcomes. We further describe the features and validation procedures in the **Method** below.

Using a pre-registered modeling pipeline (https://doi.org/10.17605/OSF.IO/YBA58), we identified a common factor for some physical illness domains (neurological, endocrine/metabolic, genitourinary, musculoskeletal), along with more complex multifactorial structures for others (circulatory, respiratory, cancer, gastrointestinal) (see Supplementary Figs. 1-8). These factor models all provided a good fit to the data as evidenced by Confirmatory Fit Indices (CFIs) > 0.9 and Standardized Root Mean Square Residuals (SRMR) < 0.1. In cases where multiple factors were identified within a domain, each latent factor represents subgroups of physical illness phenotypes that exhibit greater levels of genetic overlap compared to the other conditions within that domain. For example, in the respiratory domain, we identified a factor defined by traits with an atopic component (asthma, nasal polyps) and another factor characterized by high genetic overlap in traits with central pulmonary pathology (pneumonia, chronic airway obstruction, respiratory abnormalities). A list of the factors within each domain and the traits that define them is presented in Supplementary Table 1, with model fit indices for each domain presented in Supplementary Table 2.

### Relationships between Physical & Psychiatric Illness Latent Factors

We took forth the factor structures identified in each physical illness domain and modeled them alongside five previously established latent psychiatric factors^18^ representing the genetic overlap between distinct groups of compulsive, psychotic/thought, neurodevelopmental, internalizing, and substance use disorders (see Fig. 1). To determine the extent to which factor correlations are operating through common pathways or pathways driven by specific pairs of traits we utilized the Q_Factor_ heterogeneity index^19^. A significant Q_Factor_ statistic indicates that the inter-factor correlations fail to recapture the associations across the conditions that define the factors. For example, Q_Factor_ would likely be significant if two physical illnesses on one factor had directionally opposing relationships with the disorders that define a psychiatric factor. Q_Factor_ thereby distinguishes between broad risk-sharing patterns via the factor and risks that are more circumscribed than that implied by the factor correlation.

**Fig. 1:**
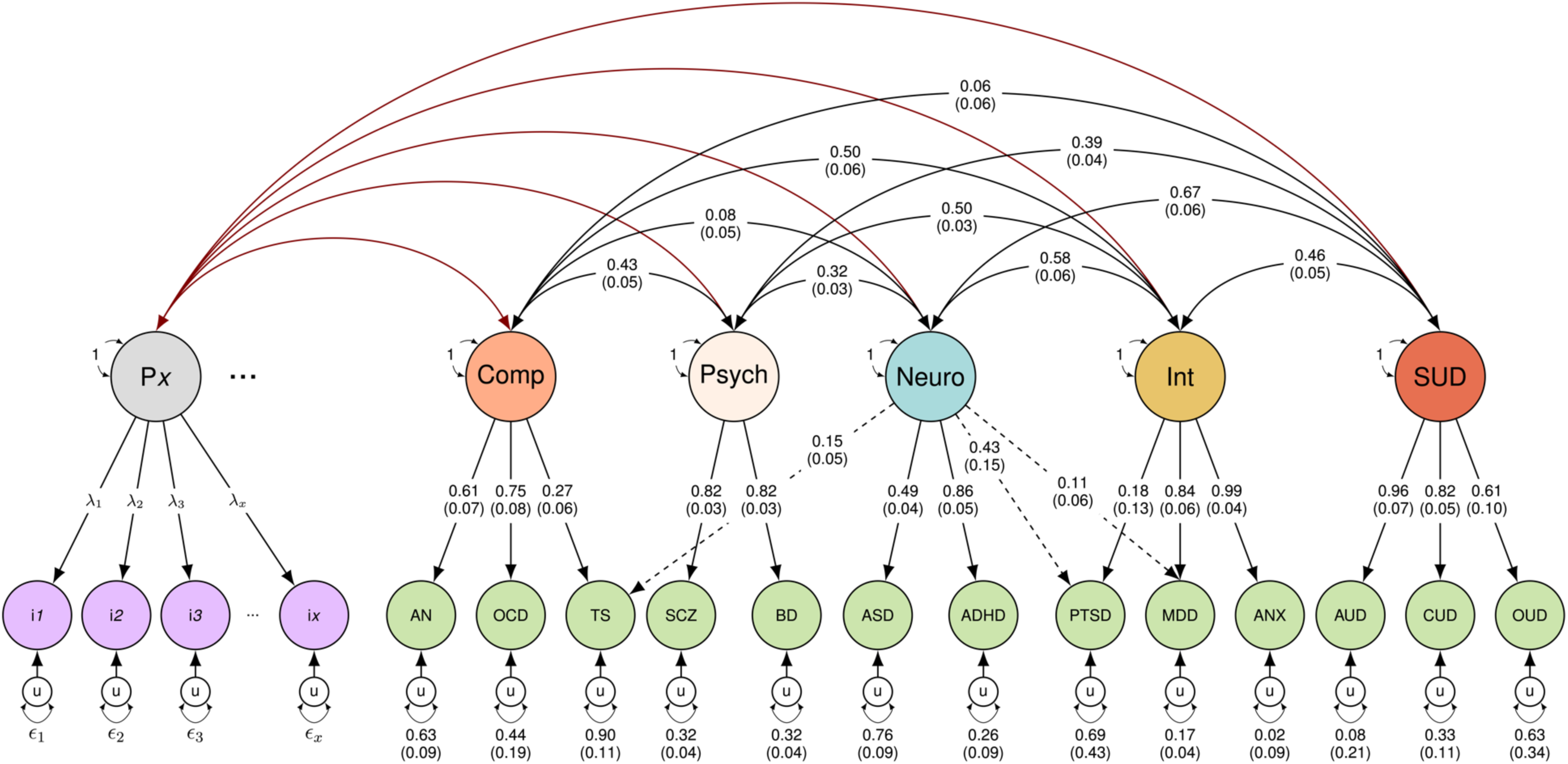
Schematic of primary analysis. *Note.* Single-headed arrows represent regression paths. Curved double-headed arrows represent correlations among the (residual) genetic variance components for each trait. Each *u* represents residual variances for the psychiatric and physical traits. The three dots next to P*x* depict a truncated factor structure for physical illness domains where a multifactorial solution was identified. Comp, Compulsive; Psych, Psychotic/Thought; Neuro, Neurodevelopmental; Int, Internalizing; SUD, Substance-use Disorders.

Across all eight physical illness domains, we identified 37 significant inter-factor correlations with latent psychiatric factors that were significant at a Bonferroni corrected threshold of *p* < 5.6 × 10^−4^ (0.05/90 inter-factor correlations) and not significant for Q_Factor_ at the same significance threshold. Fig. 2 visually depicts the inter-factor genetic correlations (*r_g_*’s) across the physical and mental illness domains, with the corresponding numeric values presented in Supplementary Table 3. Overall, our results revealed distinct patterns of relationships depending on the psychiatric domain. For the compulsive factor, we observed a protective effect with physical illness factors in the circulatory and endocrine/metabolic domains (Mean *r_g_*: -0.32; Range: -0.43 to -0.20). In the domain of psychotic/thought disorders, we see a general trend of null relationships across physical illness domains, apart from significant positive correlations with respiratory and gastrointestinal illness domains (Mean *r_g_*: 0.17; Range: 0.14 to 0.20). We further observe significant Q_Factor_ results between psychotic/thought disorders and factors in the circulatory and genitourinary domains, indicating divergent associations across the disorders within these domains that do not operate at the level of the factors. The pattern of bivariate *rg’s* between schizophrenia (SCZ), bipolar disorder (BD), and the physical illness phenotypes, revealed that these significant Q_Factor_ results were being driven by BD, which exhibited greater *rg’s* than SCZ with both circulatory and genitourinary traits.

**Fig. 2:**
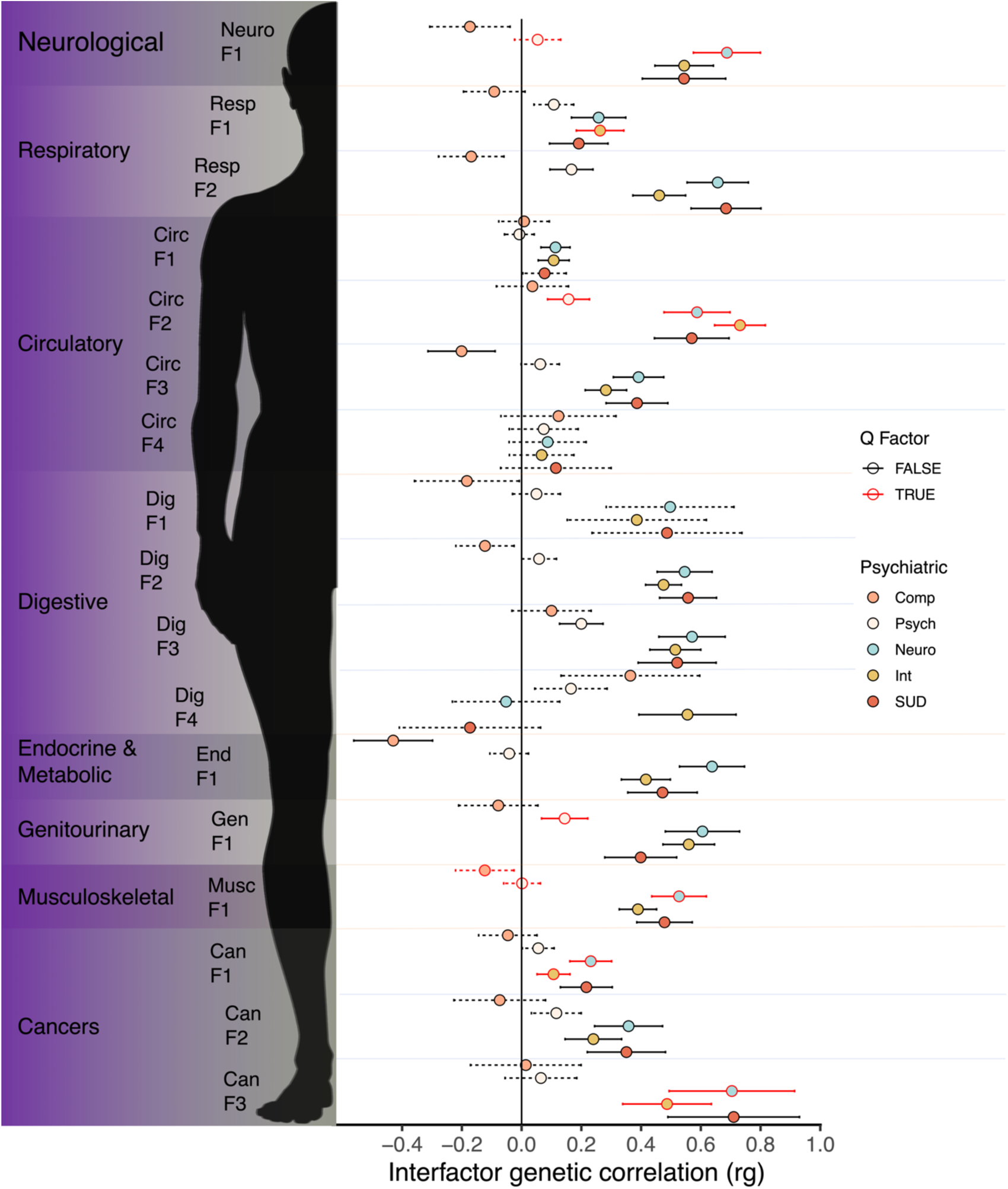
Inter-factor relationships across physical and psychiatric illness domains. *Note*. Inter-factor correlations across factor structures identified in eight physical illness domains and five psychiatric factors. Dashed bars represent traits not surpassing a Bonferroni-corrected significance threshold of *p* < 5.6 × 10^−4^ and orange bars represent significant Q_Factor_ results at the same threshold. Translucent orange lines delimit separate physical domains, with translucent blue lines delimiting separate factors within domains. Comp, Compulsive; Psych, Psychotic/Thought; Neuro, Neurodevelopmental; Int, Internalizing; SUD, Substance-use Disorders.

In contrast, we observed pervasive and positive inter-factor relationships across a wide range of factors in the physical illness domains and the neurodevelopmental, internalizing, and substance use disorder factors. Within the neurodevelopmental domain, we observed moderate to high inter-factor correlations with factors across all physical illness domains (Mean *r_g_*: 0.49; Range: 0.11 to 0.70). However, these correlations were partly driven by significantly heterogeneous illness effects, such as the relationships with the musculoskeletal and neurological domains and specific factors within the circulatory and cancer domains. Inspection of the pairwise genetic correlations between physical illnesses and the disorders that define the neurodevelopmental factor indicates that these Q_Factor_ results were driven by larger *r_g_*’s with attention deficit hyperactivity disorder (ADHD) relative to the other disorders on the neurodevelopmental factor (e.g., autism spectrum disorder). This elevated pattern of genetic risk sharing for ADHD is visually depicted in Fig. 3, which displays average pairwise *r_g_*’s between each psychiatric disorder and all physical and psychiatric traits. Three findings of note here are that: (*i*) compared to all other psychiatric disorders, ADHD has the highest average genetic correlation (*r_g_*) with physical illnesses (Mean: 0.36, SD: 0.20); (*ii*) on average, ADHD was as genetically correlated with physical disorders as it is with psychiatric disorders (Mean: 0.35, SD: 0.22); and (*iii*) perhaps most surprising was that the average genetic correlation (*r_g_*) between ADHD and physical illnesses was higher than the average genetic correlation (*r_g_*) between physical illnesses with other physical illnesses outside of their respective physical illness domain (Mean: 0.27, SD: 0.23). This pattern of results collectively indicates that the genetic signal for ADHD indexes particularly broad risk pathways that transcend psychiatric and physical illness diagnostic boundaries.

**Fig. 3:**
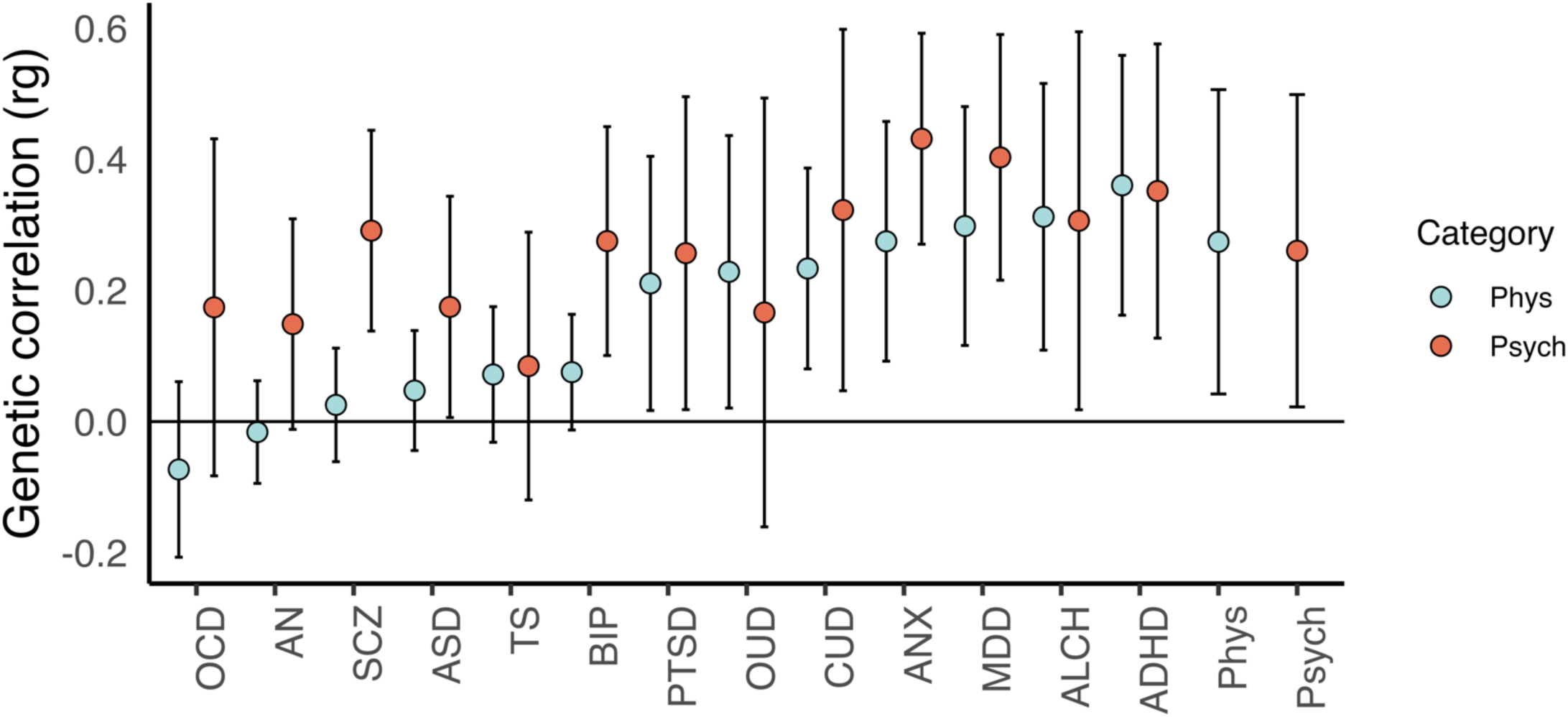
Mean pairwise relationships between 13 psychiatric and 76 physical illness traits. *Note.* Individual dots depict mean pairwise genetic correlations (*rgs)* between the 13 psychiatric traits and other psychiatric and physical traits. The error bars depict one standard deviation above and below each mean. The last two bars depict means and standard deviations (SDs) for pairwise *rgs* between all unique physical-physical and psychiatric-psychiatric combinations. For physical-physical, the mean and SD reflect average *rgs* between all physical traits and traits outside of each physical domain (e.g., mean *rg* between circulatory traits and traits in the other seven physical health domains). Phys, physical; Psych, psychiatric.

For internalizing disorders, we observed significant inter-factor correlations between at least one of the factors in all the physical illness domains (Mean *rg*: 0.41; Range: 0.11 to 0.73). However, we observed significant heterogeneous (Q_Factor_) results between the internalizing factor and factors in the respiratory, circulatory, and cancer domains. The pattern of genetic correlations for specific traits within the internalizing factor indicates that this was driven by larger genetic correlations for major depressive disorder (MDD) relative to the other internalizing disorders, with the average genetic correlation (*rg*) between MDD and physical illnesses (Mean: 0.30, SD: 0.18) being second only to ADHD. Lastly, we observed significant positive relationships between the substance use domain and factors across all physical illness domains (Mean *rg*: 0.47; Range: 0.19 to 0.71), with no significant Q_Factor_ results for any of the physical illness factors. Unlike neurodevelopmental and internalizing disorders, the absence of Q_Factor_ findings for the substance use factor indicates that the genetic signal that is shared across this group of psychiatric disorders is more broadly shared with the genetics underlying physical illness.

### Factor Analysis across all Physical Traits

The physical illness domains utilized in the primary analysis were chosen based on prior epidemiological work and reflect groupings of traits based on the body system where the cardinal symptom or disease manifests. Contained within this categorization system is the embedded assumption that disorders that share disease manifestation in a certain body system will, in turn, have a shared etiology. To evaluate this assumption, we conducted a follow-up analysis using a data-driven, system-agnostic model across all 76 curated physical traits. We began by fitting a common factor model across all physical illness measures and then pruned out any traits with a standardized factor loading cut-off of < 0.6. This yielded a common factor across 34 physical traits spanning all eight domains of illness that fit the data well (CFI = 0.90; SRMR = 0.09; see Supplementary Table 4 and Fig. 4). We then modeled the common physical factor alongside the five psychiatric factors utilized in the primary analysis, using the same Bonferroni corrected threshold (*p* < 5.6 × 10^−4^) to identify significant inter-factor correlations. Consistent with the large, pervasive genetic correlations observed within physical domains, we identified a large inter-factor correlation with the substance use factor (Fig. 5; Supplementary Table 5). In addition, we identified four significant Q_Factor_ results with the compulsive, psychotic/thought, neurodevelopmental, and internalizing factors, indicating that a common physical disease factor does not generally index common genetic risk pathways shared by groupings of psychiatric disorders.

**Fig. 4:**
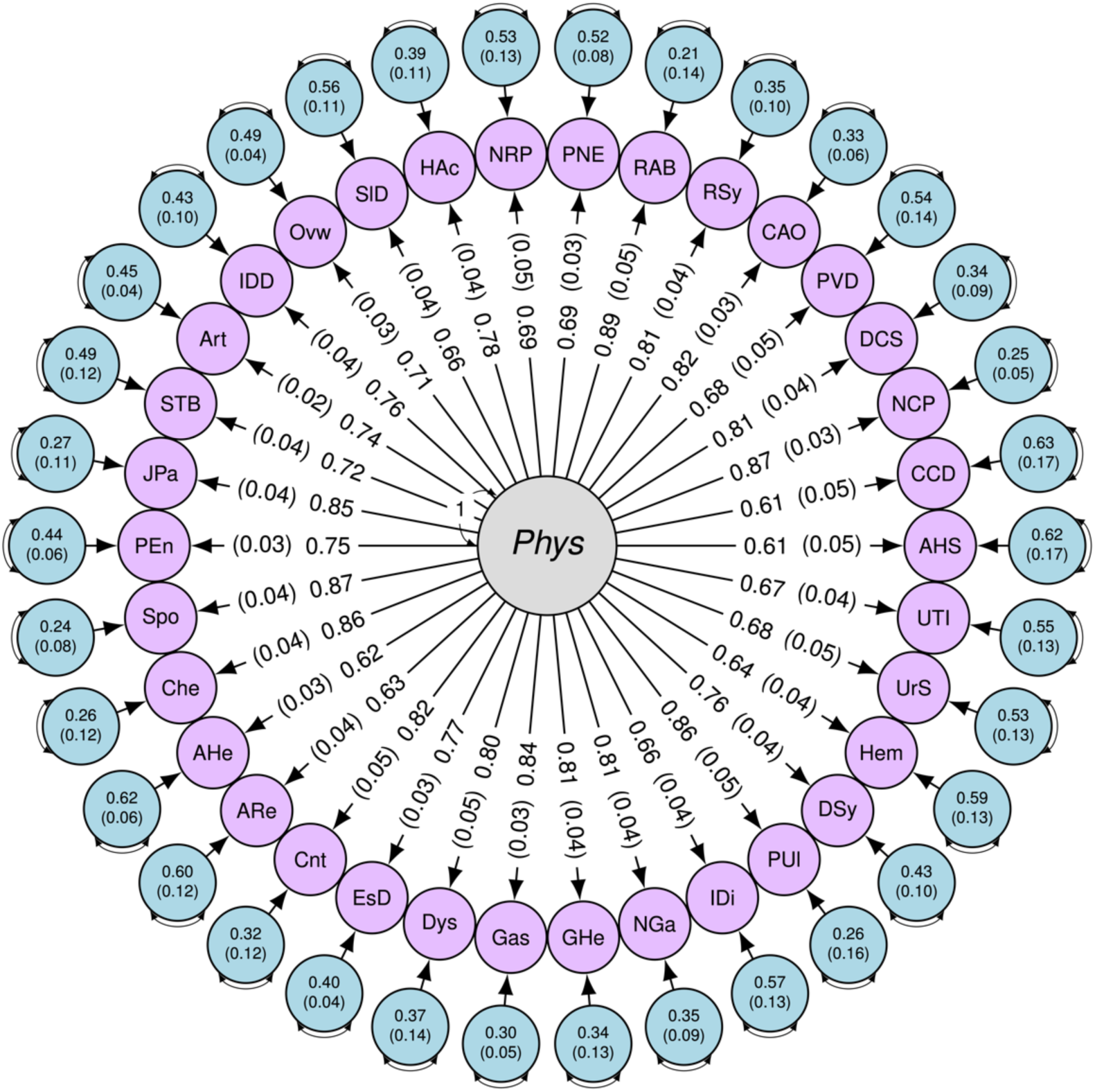
Common factor of 34 physical illness phenotypes spanning all eight body systems. *Note.* Single-headed arrows represent regression paths and curved double-headed arrows represent residual genetic variance components. The outer circles with arrows pointing to each indicator represent indicator residual variances, with the corresponding value and SE presented in each circle. AHS, abnormal heart sounds; CCD, cardiac conduction disorders; NCP, nonspecific chest pain; DCS, disorders of circulatory system-other; PVD, peripheral vascular disease; CAO, chronic airway obstruction; RSy, symptoms of respiratory system-other; RAB, respiratory abnormalities; PNE, pneumonia; NRP, nerve, root, and plexus disorders; Hac, headache syndromes; SlD, sleep disorders; Ovw, overweight, obesity, and hyperalimentation; IDD, intervertebral disc disorders; Art, arthropathies; STB, disorders of synovium, tendon, and bursa-other; JPa, joint pain; PEn, peripheral enthesopathies & allied syndromes; Spo, spondylosis & allied syndromes; Che, chemotherapy; AHe, abdominal hernia; ARe, anal & rectal conditions; Cnt, constipation; EsD, diseases of esophagus; Dys, dysphagia; Gas, gastritis & duodenitis; GHe, gastrointestinal hemorrhage; Nga, noninfectious gastroenteritis; Idi, disorders of intestine-other; Pul, pepctic ulcer excl. esophageal; DSy, Digestive system symptoms; Hem, hematuria; UrS, Symptoms & disorders of urinary system-other; UTI, urinary tract infection.

**Fig. 5:**
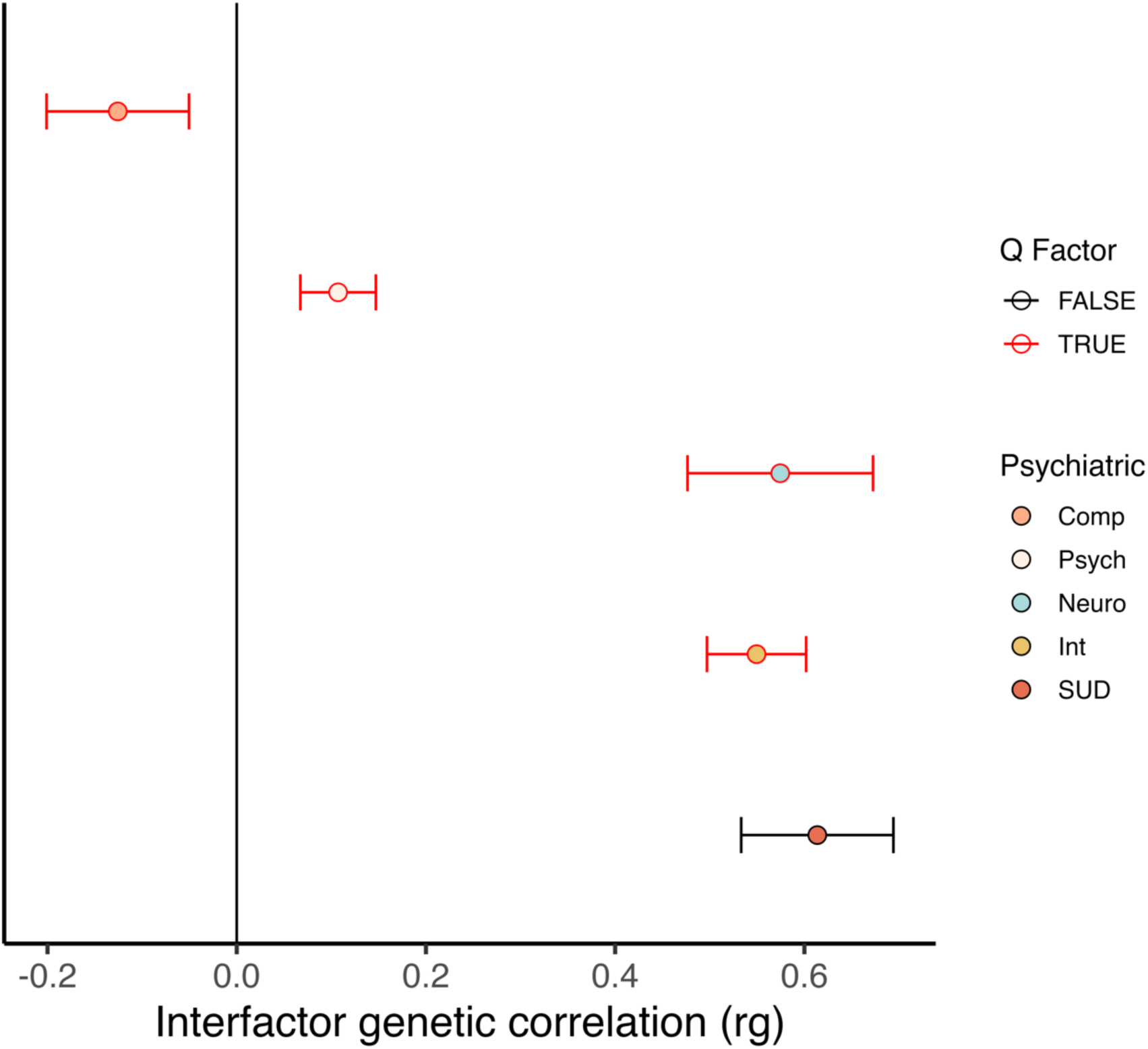
Inter-factor relationships between the common physical factor and the psychiatric factors. *Note.* Inter-factor correlations between the common disease factor and five psychiatric factors. All relationships were significant at a Bonferroni-corrected significance threshold of *p* < 5.6 × 10^−4^ with orange bars depicting significant Q_Factor_ results at the same threshold. Comp, Compulsive; Psych, Psychotic/Thought; Neuro, Neurodevelopmental; Int, Internalizing; SUD, Substance-use Disorders.

## Discussion

Prior research consistently indicates that the comorbidity problem extends beyond the psychiatric space to include physical conditions^20–24^. However, the extant epidemiological literature has primarily focused on analyzing relationships between psychiatric and physical comorbidity in single cohorts. Here we present the first multivariate genomic investigation and the most comprehensive analysis of psychiatric and physical comorbidity to date. Our findings provide a detailed characterization of broad versus specific genetic risk pathways that connect clusters of psychiatric and physical traits. Leveraging the largest publicly available GWAS summary statistics for 76 physical illness traits, we systematically modeled the complex factor structure across eight major domains of physical illness. Through further modeling with established clusters of psychiatric illness, our results reveal substantial genetic risk sharing across specific clusters of psychiatric and physical conditions. In contrast to phenotypic findings that might examine comorbidity with physical conditions in a sample ascertained for a psychiatric disorder, the current estimates are derived from genomic data for different participant samples. The fact that we still observe such pervasive genetic overlap has important implications for interpreting emergent GWAS signals, etiological models, and our current nosology.

The prior phenotypic literature for thought disorders has identified numerous physical comorbidities across multiple illness domains for both schizophrenia^25,26^ and bipolar disorder^27,28^ (e.g., respiratory, neurological, and gastrointestinal disorders), with meta-analyses indicating a comorbid physical disease among ∼50% of schizophrenia cases^29^. Our results provide mixed support for prior phenotypic findings. We find that psychotic/thought disorders have null genetic relationships with factors across the eight physical illness domains, except for positive genetic correlations with subsets of respiratory and gastrointestinal outcomes. In addition, the phenotypic literature has linked compulsive disorders to increased risk for several physical comorbidities in the metabolic, musculoskeletal, and circulatory domains^30,31^. In contrast, we observed protective associations with endocrine/metabolic and circulatory illnesses. An interpretation for this inconsistency could be that phenotypic findings are indexing shared risk factors that are not captured by the common genetic variation that was measured in the current analyses (e.g., upstream environmental risk factors or rare variants).

Epidemiological literature has documented extensive physical comorbidities across disorders in substance use, internalizing, and neurodevelopmental domains^6,21,24,32,33^. Consistent with prior findings, we identify genetic overlap between the substance use factor and factors in all eight physical illness domains. The substance use factor was also the only psychiatric domain to show significant genetic overlap with a common factor defined by 34 physical illness outcomes spanning eight body systems. This indicates that, compared to other psychiatric disorders, the genetic variance shared across multiple substance use illnesses is also shared by a particularly broad range of physical outcomes. The internalizing factor also exhibited genetic overlap with at least one factor in all eight physical illness domains. However, certain associations appeared to be more disorder-specific (e.g., specific factors within the respiratory and circulatory domains), with MDD exhibiting the greatest mean genetic correlation (*r_g_*) with physical traits among internalizing disorders. In the domain of neurodevelopmental disorders, we observed a similar pattern of broad and disorder-specific risk sharing. The pattern of bivariate genetic correlations between neurodevelopmental disorders and physical illnesses suggests that ADHD, which also displayed a higher mean genetic correlation with physical rather than psychiatric disorders, is likely driving much of the disorder-specific overlap with physical health outcomes. Although we identify relationships with physical illnesses operating via the factor for substance use disorders, the heterogeneity observed within internalizing and neurodevelopmental disorders underscores the importance of employing a nuanced statistical approach capable of examining varying levels of risk sharing.

The theoretical dichotomy between psychiatric and physical illness reflects a historical divide between conditions of functional origin (e.g., without identifiable physical changes, often assumed to be psychological in origin) and organic origin (e.g., characterized by identifiable physical changes), with psychiatry inheriting the former and other specialized medical disciplines the latter^4^. Psychiatry’s focus on diseases with ambiguous causal pathways has led to an acknowledgment of the dappled nature characterizing the causes of psychiatric illness. The current findings do not support this dichotomy: we observed that substantial genetic overlap exists both within and across all physical illness domains, in addition to considerable genetic overlap with groups of psychiatric disorders. Moreover, the average genetic correlations between ADHD, MDD, alcohol use disorder, and physical illness traits were substantial and higher than the average genetic correlation of physical with other physical illness measures outside their respective domain. This indicates that the genetic signal for physical outcomes is diffuse and highly overlapping with psychiatric risk pathways and as such the emergent genetic signal can be generally characterized as capturing genetically distal causal chains that are largely not trait specific. These results suggest a reevaluation of our diagnostic classification system that reflects the interconnectedness of risk factors across specific groupings of psychiatric and physical outcomes.

### Limitations and Future Directions

The findings presented in this study should be interpreted with consideration of several limitations, chiefly the restriction to European-only GWAS summary statistics. We hope that, as datasets from other ancestral groups become publicly available and increase in size, these analyses can be extended to a more diverse set of ancestral groups. In addition, we highlight that the current results should be interpreted as reflecting genetic architecture for shared common genetic variant liability (i.e., SNPs with a minor allele frequency > 1%). Future work utilizing whole-genome sequencing should reevaluate the patterns of genetic risk sharing in the context of rare genetic variation. Finally, we note that the domain-specific and common factor structures reported above reflect clusters of genetic overlap across common physical conditions. The inclusion of rare or more specific disorders may result in alterations to the conformity of the identified physical factor structures and their relationships with groups of psychiatric illness.

The pattern of relationships across groups of psychiatric and physical disorders reported above may reflect some combination of vertical and horizontal pleiotropy. Vertical pleiotropy refers to a causal chain where a genetic variant influences a first trait which, in turn, influences a second trait. In contrast, horizontal pleiotropy reflects genetic variants that jointly influence multiple traits. Future work that disentangles these mechanisms is vital for informing how the current findings are integrated into a clinical setting. For example, identifying vertical pleiotropic effects would indicate that treatment of one disorder would have cascading effects on subsequent physical outcomes. If we take substance use disorders as an example, in line with a vertical mechanism, it could be that the use of substances has wide-ranging consequences that increase the risk for virtually all surveyed physical conditions. Alternatively, the observed patterns of genetic risk sharing may reflect horizontal pleiotropy via upstream processes that may reflect useful treatment targets for multimorbid presentations. Indeed, a horizontal explanation is just as easily generated for the substance use factor wherein some upstream process (e.g., genetically mediated personality characteristics) has far-reaching consequences across a range of psychiatric and physical outcomes.

### Conclusion

Comorbidity research across psychiatric and physical outcomes has historically been confined to describing relationships across sets of disorders in a single cohort. This research has revealed widespread overlap within and across psychiatric and physical disorders. Through the application of Genomic SEM and E-SEM, the current study brings together a wide-ranging set of physical and psychiatric disorders into a multivariate framework. Our findings uncover broad genetic liabilities spanning multiple physical and psychiatric domains along with more circumscribed risk sharing, such as the especially high overlap between ADHD, MDD, and physical illness traits. This study enriches our knowledge of the multivariate systems of relationships underlying psychiatric and physical comorbidity with implications for interpreting emergent genetic signals and reconsidering the theoretical dichotomy between these two sets of disorders.

## Method

### Trait Selection

A prior large-scale epidemiological investigation of psychiatric and physical comorbidity utilized diagnostic bins encompassing a wide range of physical conditions available in Danish national registries^6^. Building upon this, we curated an initial list of physical traits by intersecting the diagnostic bins from the prior study with the diagnostic categories described in the UK Biobank (UKB). Among the nine diagnostic categories, the hematopoietic domain did not contain sufficient traits passing the quality control (QC) filters outlined below. As a result, this domain was excluded from further modeling. To focus on more common physical conditions that frequently coexist in the population, we restricted this list to traits with over 2,000 cases in the UKB. The initial list included both the broadest category (e.g., heart and valve disorders) for each physical trait and the sub-trait within that category (e.g., aortic aneurysm). Leveraging Genomic SEM’s capability to integrate traits from multiple samples, we updated this list by incorporating the most recent GWAS summary statistics for each trait from the GWAS Catalog (a comprehensive database of genomic studies - https://www.ebi.ac.uk/gwas/) for either (a) the broad disorder category obtained in the UKB or (b) a specific disorder within the broader category, depending on which had more cases. The selection criterion was based on the number of cases rather than the total sample size due to the disproportionate number of controls for some traits. A comprehensive list of the disorders included in the modeling procedure is presented in Supplementary Table 6.

The psychiatric factors utilized in the primary analysis were selected based on a previously established factor structure^15^, which contains five correlated genomic factors. These factors, encompassing compulsive, psychotic/thought, neurodevelopmental, internalizing, and substance use disorders, represent groupings of shared genetic liability across 13 major psychiatric disorders. A detailed list of the disorders that define each psychiatric factor is presented in Supplementary Table 7.

### Genomic SEM

A standard set of QC filters was applied to all GWAS summary statistics. This set of filters was applied using the *munge* function available within Genomic SEM. Munging GWAS summary statistics involves reducing the number of SNPs to HapMap3 SNPs and then pruning by minor allele frequency (MAF) less than 0.01 and an imputation score (INFO) below 0.9 when this information is available in the summary statistics. The “munged” summary statistics were then used as input for the multivariable version of LDSC implemented in Genomic SEM. Linkage disequilibrium (LD) weights employed to estimate the LDSC regression model were obtained from the 1000 Genomes Phase 3 European LD Scores. These weights exclude the major histocompatibility complex (MHC) due to the intricate LD structure in this region that may bias estimates. Following the guidelines set by the original LDSC developers for producing interpretable genetic covariance estimates, physical traits with SNP-based heritability (*h^2^_SNP_*) *Z-* statistics < 4 were removed.

LDSC produces two meaningful sets of output: the genetic covariance and sampling covariance matrix. The genetic covariance matrix estimated with LDSC is comprised of SNP-based heritability estimates on the diagonal and genetic covariances on the off-diagonal. As these disorders are all binary outcomes, these estimates were converted to the liability scale, using the corresponding population prevalence from the original publication and, for meta-analyzed GWAS summary statistics, the sum of effective sample sizes across contributing cohorts^34^. The sampling covariance matrix comprised of squared standard errors on the diagonal (e.g., the sampling variances) and sampling covariances on the off diagonal, which reflect sampling dependencies that emerge when participant samples overlap across the included traits. The sampling covariance matrix is estimated directly from the GWAS data and is what allows traits with varying and unknown degrees of participant sample overlap to be included with appropriate parameter estimates and standard errors.

### Modeling Procedure

For each physical illness domain, we began by fitting a common factor in confirmatory factor analysis (CFA) to identify if this simple factor structure adequately recaptures the observed genetic covariance between the traits within each physical illness domain. If the common factor provided adequate model fit (as determined by CFI > 0.9 and SRMR < 0.1) then this parsimonious, single factor model was carried forward for that physical illness domain. If a common factor did not provide adequate fit, an exploratory factor analysis (EFA) was conducted using Genomic E-SEM to examine multifactorial solutions, a novel extension that we describe and validate via extensive simulations in a separate section below. The number of factors in each EFA was guided by the optimal coordinates, acceleration factor^35^, and Kaiser rule^36^ calculated from covariance matrices from each physical illness domain that were not adequately accounted for by a common factor. Next, we took forth traits in the EFA with standardized factor loadings exceeding our preregistered cutoff of 0.4 to be modeled in a subsequent CFA.

We assessed the associations between each physical illness factor and psychiatric factor by modeling each physical illness domain model alongside five previously established psychiatric factors^18^. The inter-factor correlations within this model represent the degree to which shared signal captured by the physical and psychiatric factors is also shared across factors. However, these factor correlations may not always do a sufficient job of describing the pairwise genetic associations of the disorders across factors. To evaluate heterogeneity in the inter-factor correlations we then utilized Q_Factor_ for each physical and psychiatric factor correlation. Q_Factor_ is a heterogeneity metric that becomes more significant when the factor correlation is unable to recapture the individual associations across disorders. For example, if two disorders loading on one factor have directionally discordant associations with the disorders loading on another factor then a single factor correlation will be insufficient for describing this divergent pattern of relationships. Q_Factor_ is a χ^2^ distributed test statistic, with degrees of freedom equal to one less than the cross-factor indicator correlations. Pairwise associations were also calculated to follow up on Q_Factor_ results. We first calculated the mean bivariate genetic correlation (*r_g_*) between each of the 13 psychiatric disorders and all other psychiatric and physical conditions. Next, we calculated the mean *r_g_* amongst all psychiatric and physical disorders, while removing redundant values, with the mean *r_g_* amongst physical disorders calculated between traits outside each of their respective domains (e.g., *r_g_* between neurological outcomes and traits in the other seven physical domains).

### Genomic E-SEM

*Overview.* Genomic Exploratory Structural Equation Modeling (E-SEM) is a robust framework for identifying the genomic factor structure among a set of variables with limited or no *a priori* structure. This method allows for the identification of latent variables from genomic data without predefining the entire factor structure. Moreover, Genomic E-SEM allows for the integration of subsets of traits that possess a predefined factor structure. Genomic E-SEM improves upon existing software for conducting genomic EFAs by implementing a weighted least squares (WLS) estimator. The WLS estimator prioritizes reducing misfit in cells of the genetic covariance matrix that are more precisely estimated (e.g., have a smaller *SE*). Although a WLS estimator will produce reliable model parameter estimates when the model is identified, the ‘naïve’ standard errors (*SEs*) and fit statistics generated in stage 2 estimation will be invalid, due to neither estimate utilizing the full sampling covariance matrix (*V_S_*)^16^. Therefore, robust correction techniques are necessary to obtain valid *SEs* and model fit statistics. We obtain the correct sampling covariance matrix (*V*_θ_) in stage 2 estimation through the application of a sandwich correction^37,38^:

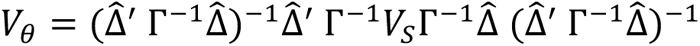

where 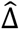 is a matrix of model derivatives at each parameter estimate, and Γ is the ‘naïve’ weights matrix estimated in stage 2 prior to correction.

*Simulations*. Key parameters of Genomic E-SEM were validated using simulated LDSC objects for 10% heritable phenotypes with a univariate LDSC intercept of 1.05 to model slight levels of population stratification. These simulations validated the sandwich-corrected *SEs* and determined the power to discern different population structures in an EFA conducted with Genomic E-SEM. For each model, we simulated participant samples of 50,000 or 500,000. The sandwich-corrected *SEs* were validated on the simulated population sizes of 50,000 and the power to discern different population structures was calculated on both sets of simulated population sizes. The first population generating structure was a two-factor model, where each factor had three indicators with a standardized loading of 0.71. We ran three iterations of this model using inter-factor correlations of 0.3, 0.5, and 0.7, with 100 simulations per iteration (e.g., 300 simulations). The second was a common factor generating model using standardized factor loadings across all indicators of either 0.32, 0.5, or 0.71 with 100 simulations per iteration (e.g., 300 simulations). All phenotypes were specified to be 10% heritable in the population. The power to detect the true population structures across the population-generating models is presented in Supplementary Fig. 9, with the validation of sandwich-corrected *SEs* presented in Supplementary Fig. 10. Across simulations, we find valid sandwich-corrected *SE*s, with ratios of sandwich corrected *SEs* to empirical *SEs* (calculated as the standard deviation of the parameter estimates across the 100 simulations) close to 1. In addition, we find adequate power to discern population structures, with power scaling in the expected direction. That is, we observed greater power to detect the true factor structure as factor loadings increased in the case of a common factor generating model and as inter-factor correlations increased in the case of a two-factor generating model. At a simulated population size of 50,000 power was ≥ 78% when the standardized inter-factor correlation was 0.5 in a two-factor generating model and when the standardized factor loading was 0.71 (∼50% of the variance explained by the factor) in a common-factor generating model. When the simulated population size reached 500,000 power was ≥ 99% across all generating structures.

### Factor Analysis across all Physical Illness Traits

The physical illness domains were chosen based on prior epidemiological work on psychiatric and physical comorbidity^6^. These domains largely reflect physical illness grouped by the body system where the cardinal symptom or disease manifests, with the implicit assumption that these disorders will, in turn, have a shared etiology. Given prior associations with psychiatric illness observed in the phenotypic space using these illness domains, we retained these broad system-based categories for the primary analysis to produce a set of results that are comparable to extant phenotypic literature. In addition, we also conducted a follow-up analysis to explore the factor structure across all curated physical illness traits (e.g., largest publicly available GWAS; *h^2^_SNP_ Z*-statistic > 4) and subsequent relationships with the five psychiatric factors utilized in the primary analysis.

Our data-driven follow-up analysis was conducted in multiple stages. We first used the acceleration factor, optimal coordinates, and Kaiser rule to determine the number of factors in the EFA. Each of these indices suggested a different number of factors, so we opted for the most parsimonious factor structure, which was a common factor suggested by the acceleration factor. Next, we conducted a CFA across 76 physical illness traits in Genomic E-SEM and employed a standardized loading cutoff of 0.6 to determine which traits would be taken forward for further modeling. We then fit two subsequent CFAs to (*i*) determine model fit for our data-driven common physical illness factor and (*ii*) evaluate the relationship between a common physical illness factor and the five psychiatric factors used in the primary analysis.

## Data Availability

The data sources for each of the GWAS summary statistics are available in Supplementary Table 6 for physical phenotypes and Supplementary Table 7 for psychiatric phenotypes. No new data were collected for this project and all data utilized in this study were from publicly available sources.

## Code Availability

Genomic SEM and E-SEM are publicly available software and can be accessed at https://github.com/GenomicSEM/GenomicSEM.

## Acknowledgments

ADG, LSS, and JML are supported by NIH Grant R01MH120219. ADG and IFF are supported by NIA Grant RF1AG073593. T.T.M. is supported by NIH grant K08MH135343. SB is supported by the Shurl and Kay Curci Foundation.

