## Supplementary Figures for "Shared Genetic Liability across Systems of Psychiatric and Physical Illness"

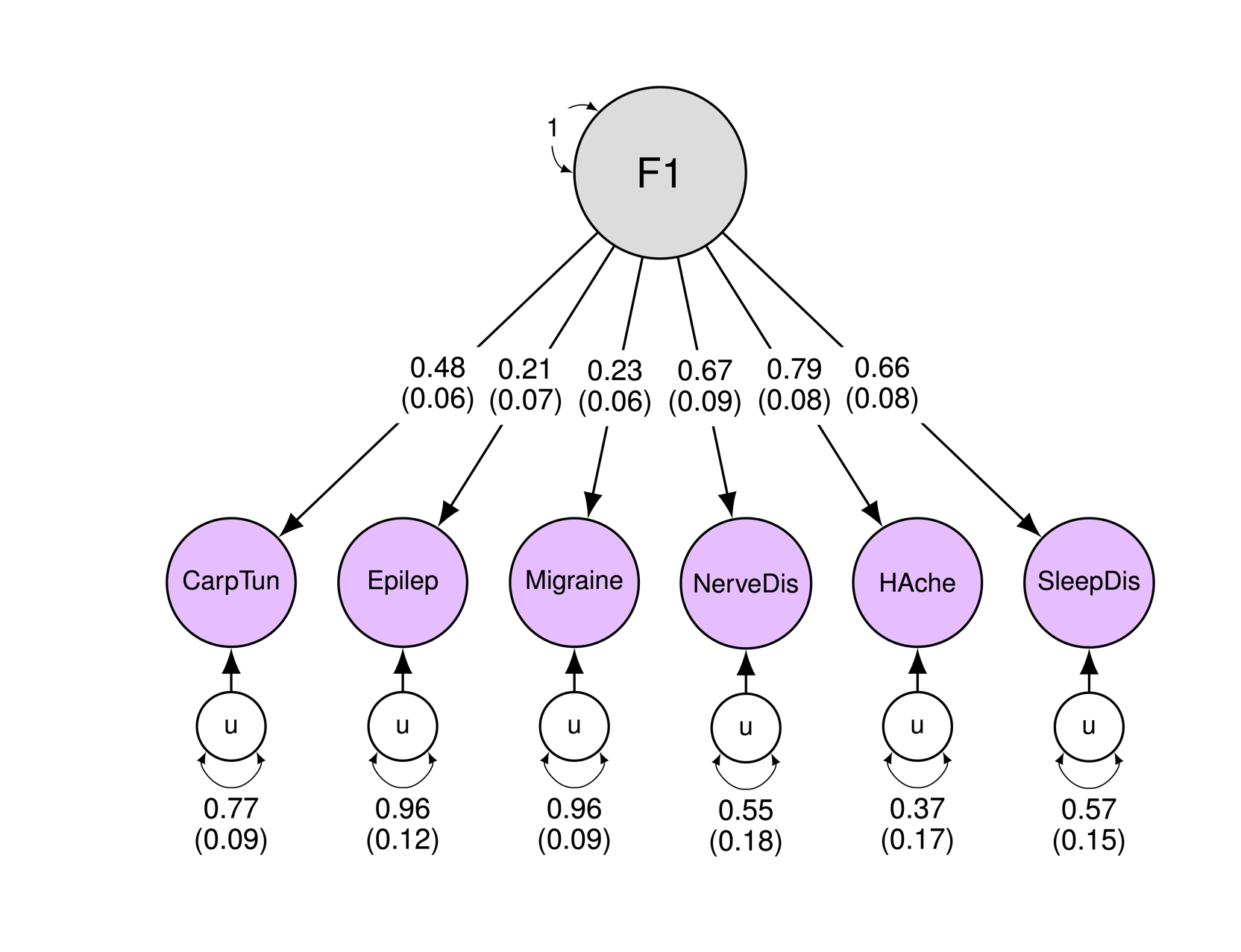
**Supplementary Fig. 1.** Path diagram of neurological domain

*Note.* Single-headed arrows represent regression paths. Curved double-headed arrows represent correlations among the (residual) genetic variance components for each trait. Each u represents residual variances for physical traits. CarpTun, Carpal tunnel syndrome; Epilep, Epilepsy; NerveDis, Nerve root and plexus disorders; HAche, Headache syndromes-other; SleepDis, Sleep disorders.

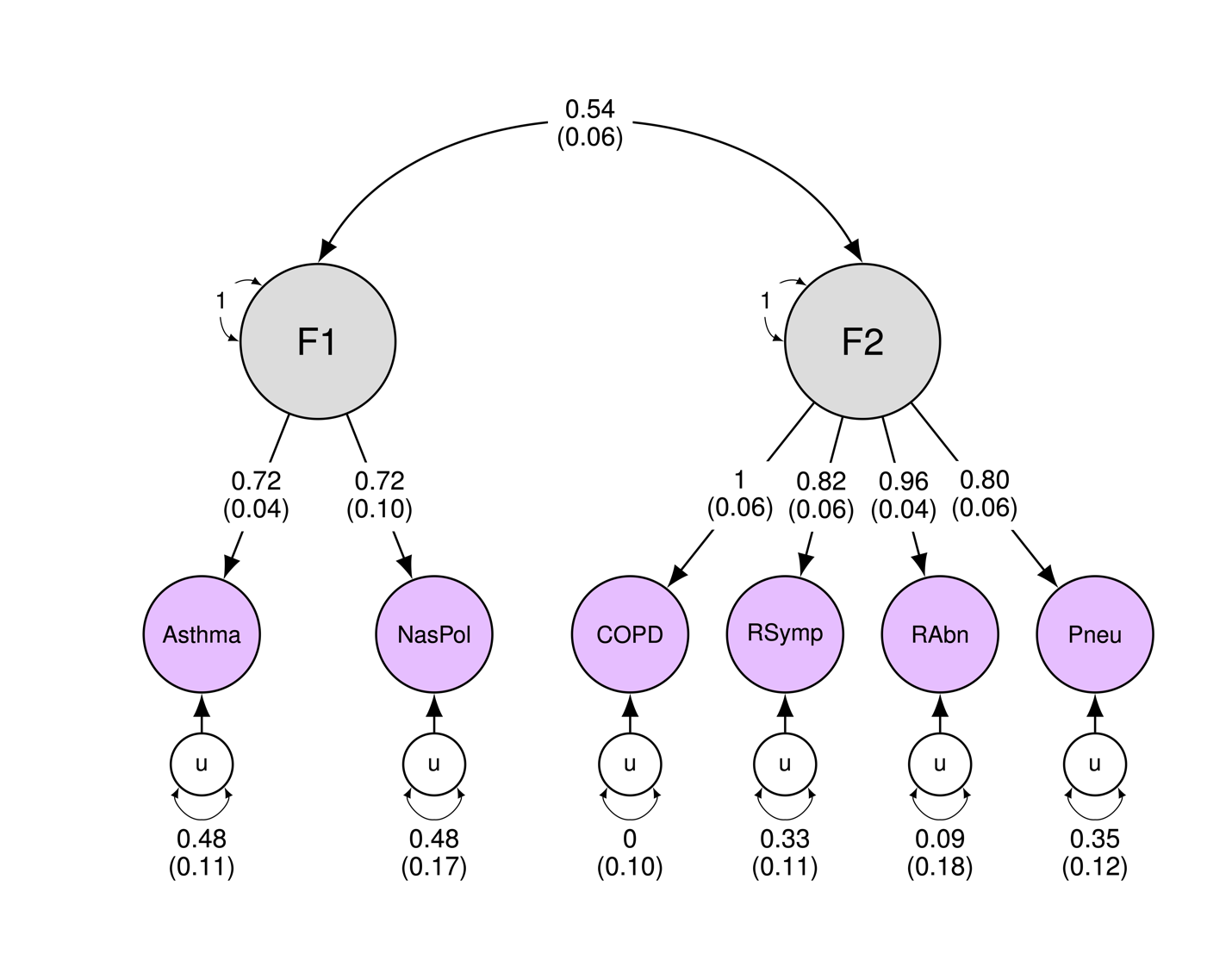
**Supplementary Fig. 2.** Path diagram of respiratory domain

*Note.* Single-headed arrows represent regression paths. Curved double-headed arrows represent correlations among the (residual) genetic variance components for each trait. Each u represents residual variances for physical traits. NasPol, Nasal polyps; COPD, Chronic airway obstruction; RSymp, Symptoms of respiratory system-other; RAbn, Respiratory abnormalities; Pneu, Pneumonia.

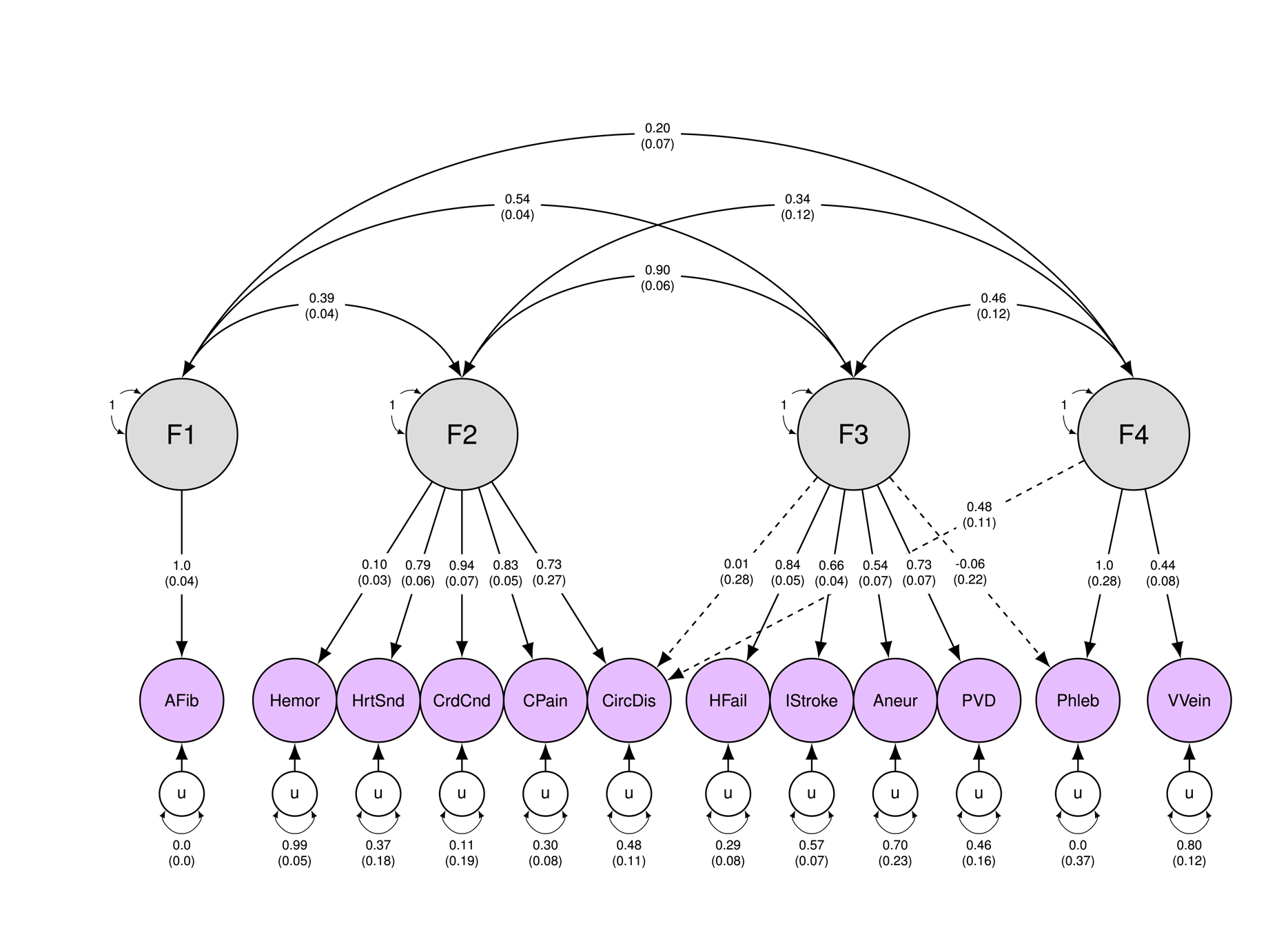
**Supplementary Fig. 3.** Path diagram of circulatory domain

*Note.* Single-headed arrows represent regression paths. Curved double-headed arrows represent correlations among the (residual) genetic variance components for each trait. Each u represents residual variances for physical traits. AFib, Atrial fibrillation and flutter; Hemor, Haemorrhoidal disease; HrtSnd, Abnormal heart sounds; CrdCnd, Cardiac conduction disorders; CPain, Nonspecific chest pain; CircDis, Disorders of circulatory system-other; HFail, Heart failure; IStroke, Cerebral ischemia; Aneur, aneurysm-other; PVD, Peripheral vascular disease; Phleb, Phlebitis and thrombophlebitis; VVein, Varicose veins.

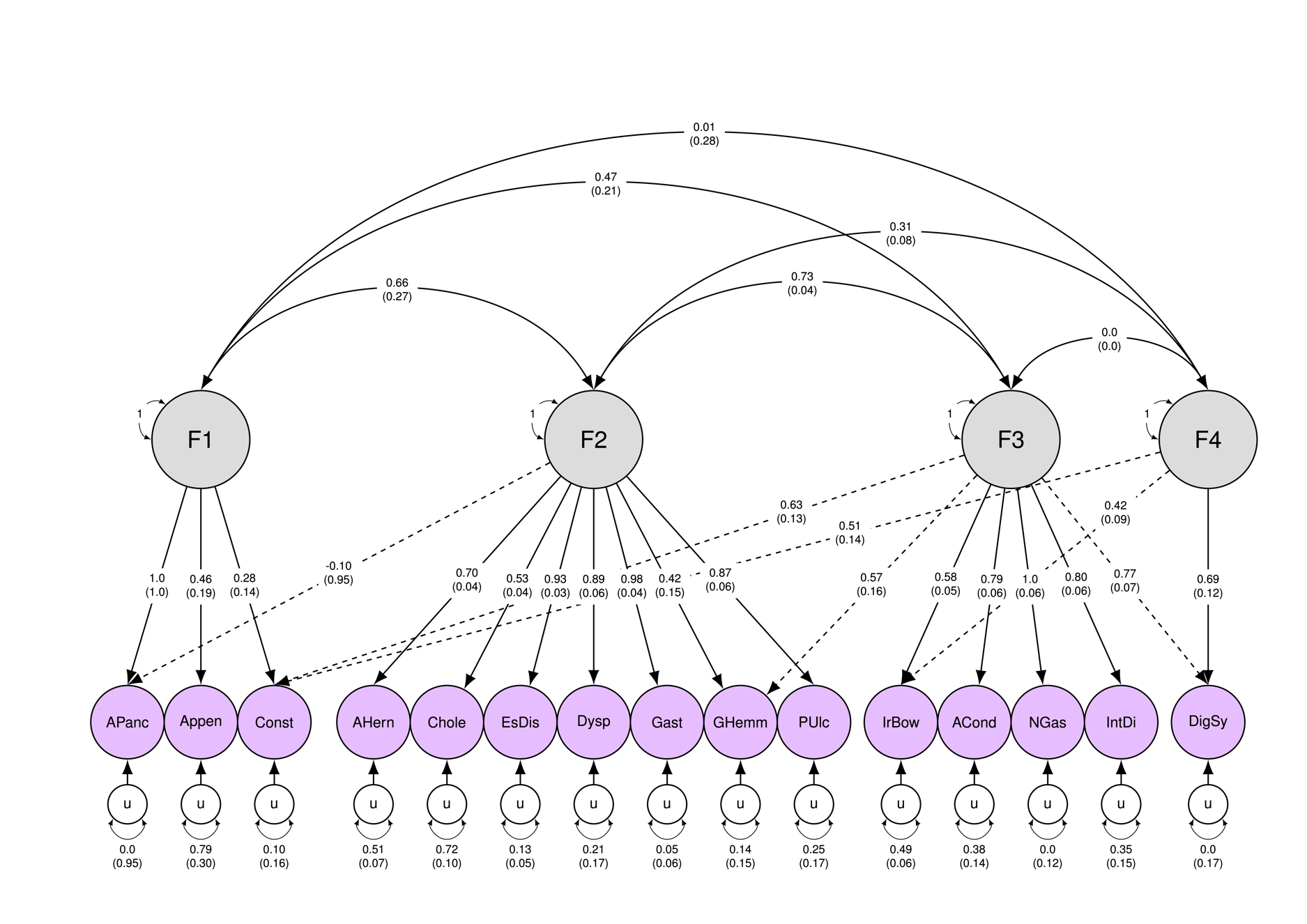
**Supplementary Fig. 4.** Path diagram of digestive domain

*Note.* Single-headed arrows represent regression paths. Curved double-headed arrows represent correlations among the (residual) genetic variance components for each trait. Each u represents residual variances for physical traits. APanc, Acute pancreatitis; Appen, Appendicitis; Const, Constipation; AHern, Abdominal hernia; Chole, Cholelithiasis and cholecystitis; EsDis, Diseases of esophagus; Dysp, Dysphagia; Gast, Gastritis and duodenitis; GHemm, Gastrointestinal hemorrhage; PUlc, Peptic ulcer excl. esophageal; IrBow Irritable bowel syndrome; ACond, Anal and rectal conditions; NGas, Noninfectious gastroenteritis; IntDi, Disorders of intestine-other; DigSy, Symptoms involving digestive system.

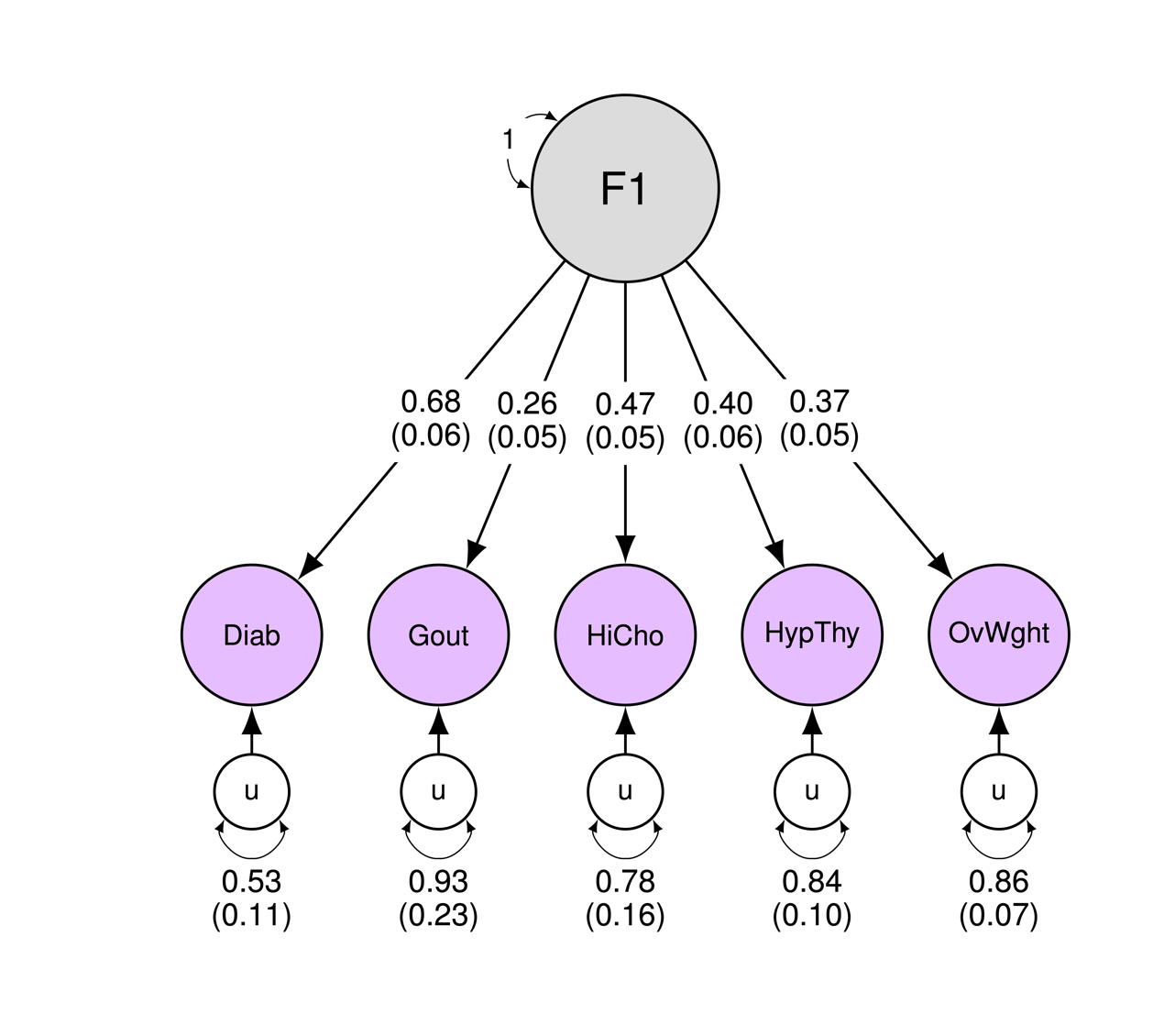
**Supplementary Fig. 5.** Path diagram of endocrine/metabolic domain

*Note.* Single-headed arrows represent regression paths. Curved double-headed arrows represent correlations among the (residual) genetic variance components for each trait. Each u represents residual variances for physical traits. Diab, Diabetes; HiCho, Hypercholesterolemia; HypThy, Hypothyroidism; OvWght Overweight, obesity, and other hyperalimentation.

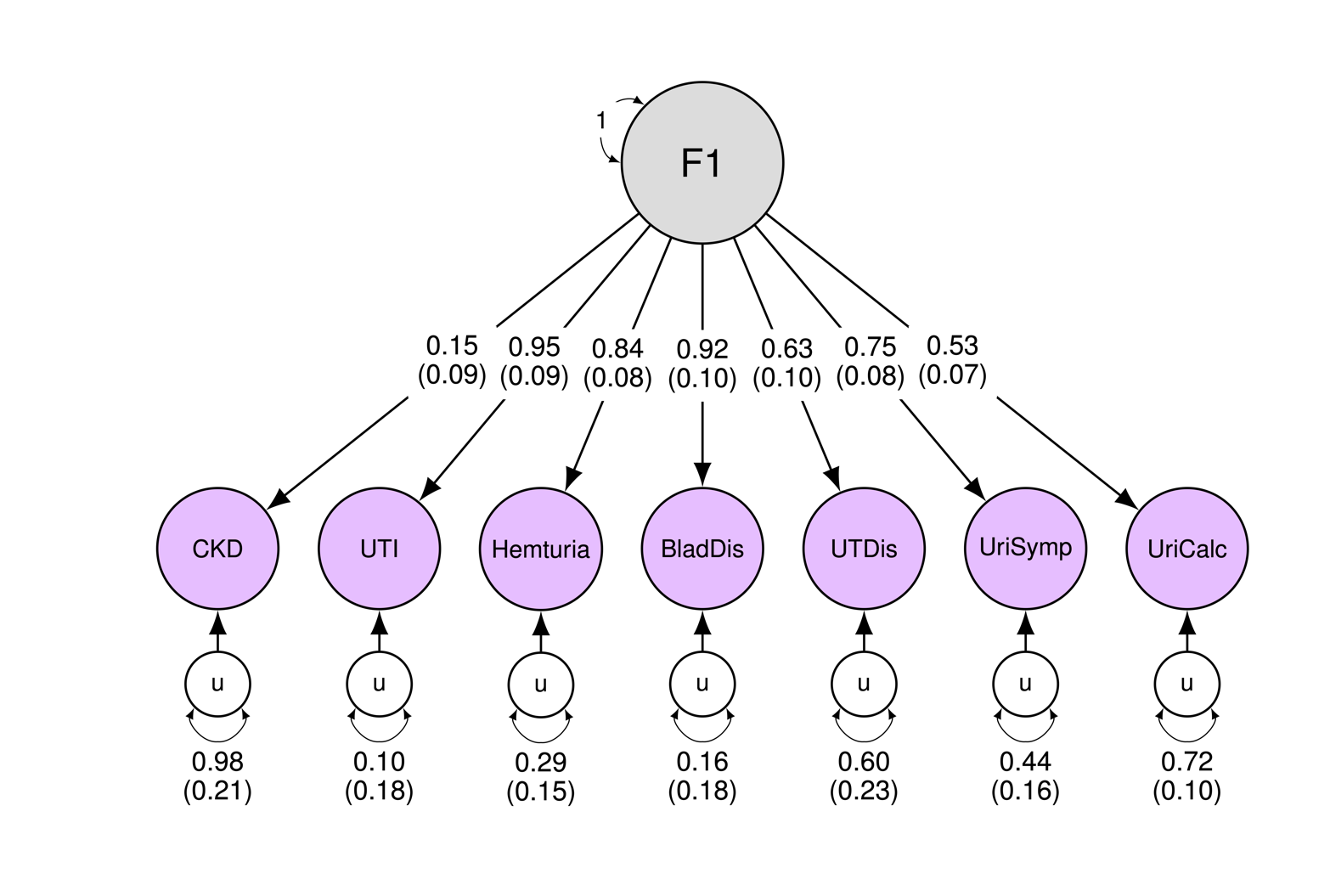
**Supplementary Fig. 6.** Path diagram of genitourinary domain

*Note.* Single-headed arrows represent regression paths. Curved double-headed arrows represent correlations among the (residual) genetic variance components for each trait. Each u represents residual variances for physical traits. CKD, Chronic kidney disease; UTI, Urinary tract infection; Hemturia, Hematuria; BladDis, Disorders of bladder; UTDis, Disorders of urethra and urinary tract-other; UriSymp, Other symptoms & disorders of the urinary system; UriCalc, Urinary calculus.

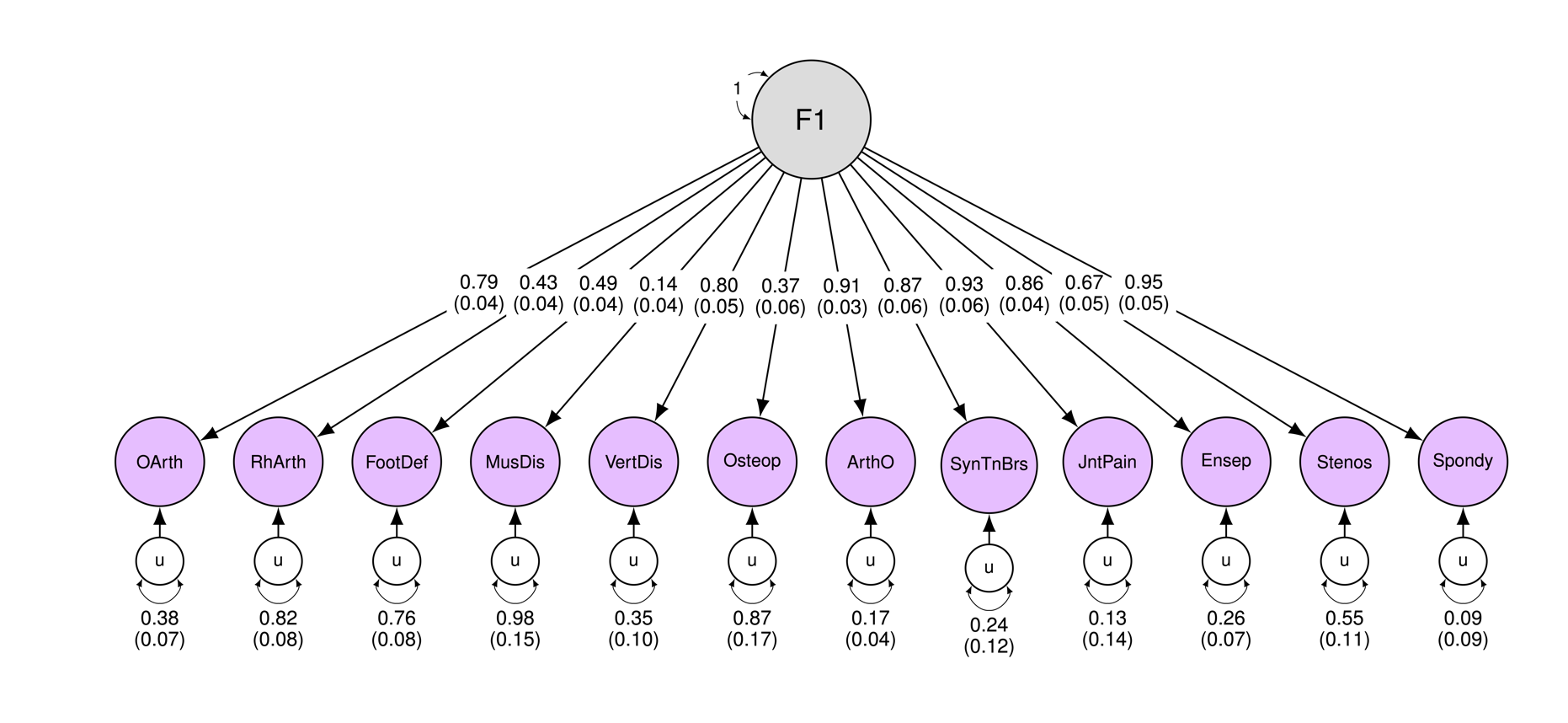
**Supplementary Fig. 7.** Path diagram of musculoskeletal domain

*Note.* Single-headed arrows represent regression paths. Curved double-headed arrows represent correlations among the (residual) genetic variance components for each trait. Each u represents residual variances for physical traits. OArth, Osteoarthritis; RhArth, Rheumatoid arthritis; FootDef, Acquired foot deformities; MuscDis, Disorders of muscle, ligament, and fascia; VertDis, Intervertebral disc disorders; Osteop, ArthO, Arthropathies-other; SynTnBrs, Disorders of synovium, tendon, and bursa-other; JntPain, Pain in joint; Ensep, Peripheral enthesopathies and allied syndromes; Stenos, Spinal stenosis; Spondy, Spondylosis and allied disorders.

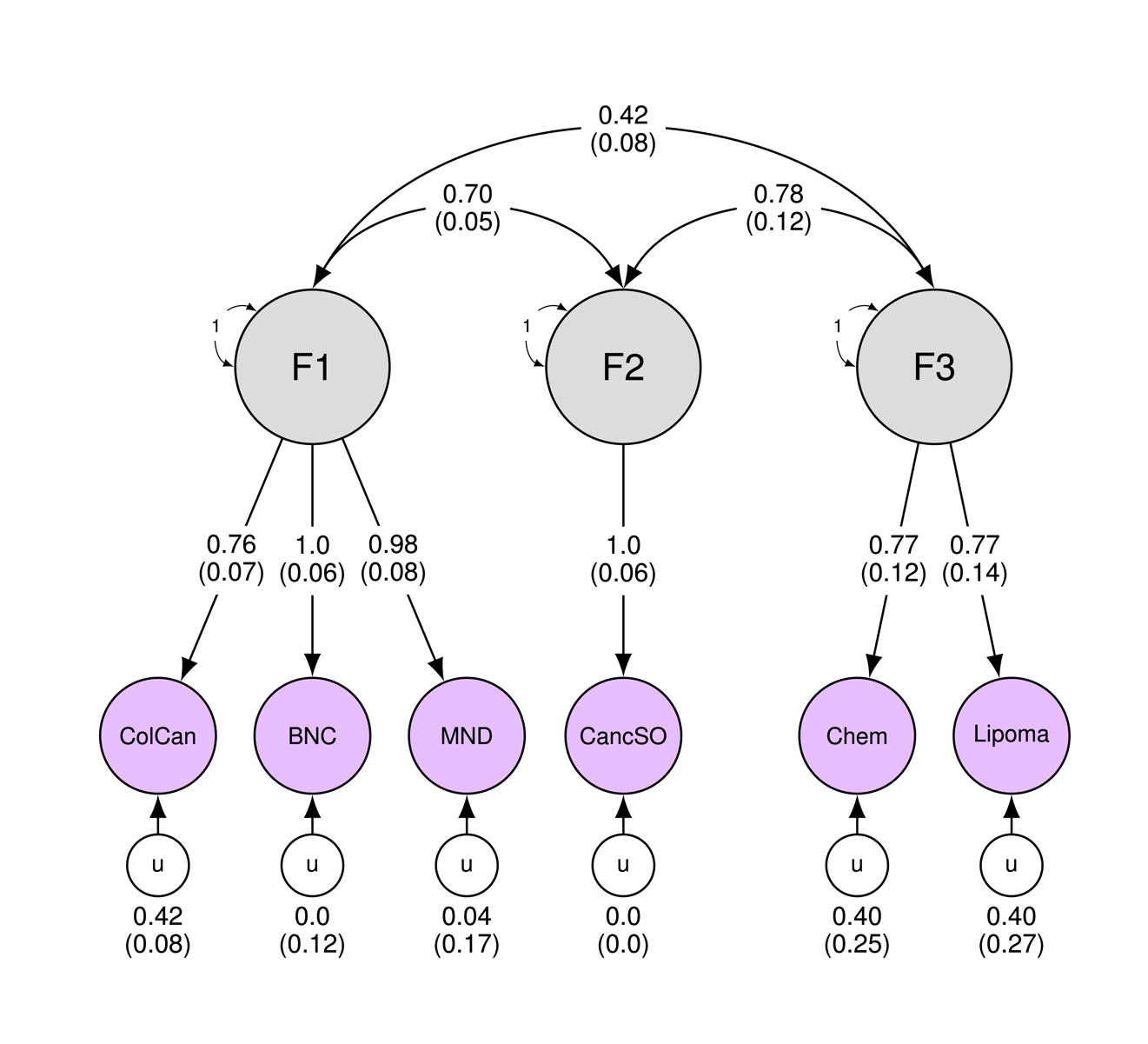
**Supplementary Fig. 8.** Path diagram of cancer domain

*Note.* Single-headed arrows represent regression paths. Curved double-headed arrows represent correlations among the (residual) genetic variance components for each trait. Each u represents residual variances for physical traits. ColCan, Colorectal cancer; BNC, Benign neoplasm of colon; MND, Malignant neoplasm of sites within the digestive organs and peritoneum; CancSO, Cancer suspected or other; Chem, Chemotherapy.

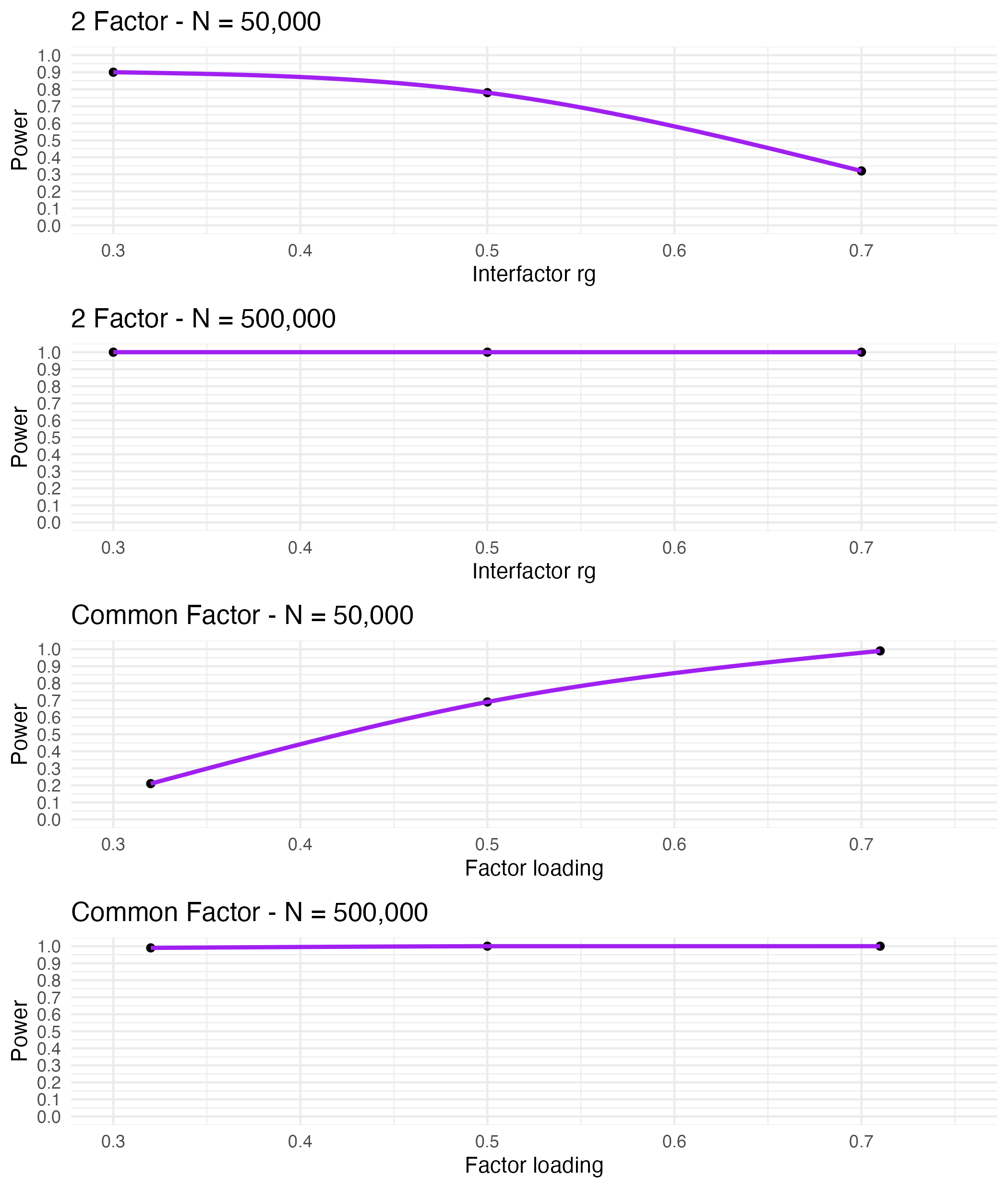
**Supplementary Fig. 9.** Power to detect population structures

**DdD**

**C**

**B**

**A**

*Note.* Power to detect true population structure across a variety of population generating models using a standardized loading cutoff 10% less than the population-generating loading. Trendlines were fit using natural cubic spindles. (A, B) Results from two-factor model with simulated populations of 50,000 and 500,000 respectively. (C, D) Results from common factor model with simulated populations of 50,000 and 500,000 respectively.

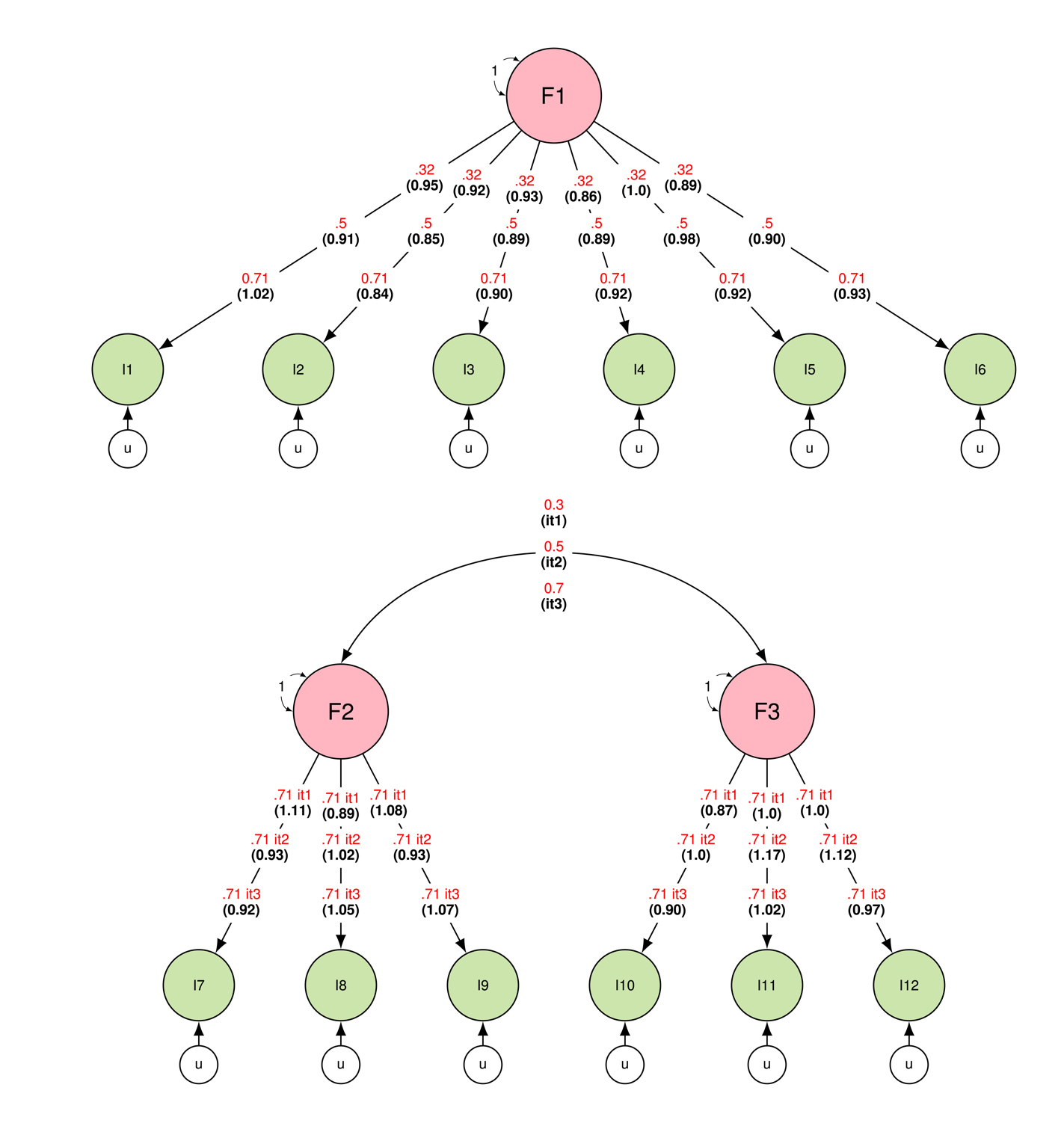
**Supplementary Fig. 10.** Genomic E-SEM sandwich-corrected standard error validation

**B**

**A**

*Note.* Single-headed arrows represent regression paths. Curved double-headed arrows represent correlations among the (residual) genetic variance components for each trait. Each u represents a residual variance. In parentheses, we provide the ratio of mean *SE* over 100 runs to the empirical *SE* (e.g., *SD* of the parameter estimates over 100 iterations) for the standardized estimates. The ratios of mean *SE* to empirical *SE* were mostly close to 1. (A) Results from the common-factor model with a simulated population of 50,000. (B) Results from the two-factor generating model with a simulated population of 50,000.
